# Diametrics: A User-Friendly Web Tool for Custom Analysis of Continuous Glucose Monitoring Data

**DOI:** 10.1101/2024.06.20.24309152

**Authors:** Catherine Russon, Michael Allen, Michael Saunby, Richard Pulsford, Neil Vaughan, Matthew Cocks, Jonathan Low, Katie Hesketh, Robert Andrews

**Affiliations:** University of Exeter Medical School; Research Software Engineering, University of Exeter; Public Health and Sport Science, University of Exeter; School of Sport and Exercise Sciences, Liverpool John Moores University; School of Health and Exercise Science, University of British Columbia - Okanagan; School of Sport, Exercise and Rehabilitation Sciences, University of Birmingham

**Keywords:** Continuous Glucose Monitoring (CGM), Web Application, Glycemic Analysis, Event-specific Analysis, Metrics, Open-source

## Abstract

**Background:** Continuous Glucose Monitoring (CGM) systems have revolutionized diabetes management by providing real-time blood glucose tracking. However, there is a need for openly accessible tools that can analyze CGM data in relation to specific events like meals or exercise, which often require extensive technical skills to interpret, thus restricting its broader use among researchers and clinicians. Developing user-friendly web applications to facilitate this analysis could significantly broaden accessibility and utility.

**Method:** *Diametrics* was built with a focus on ease-of-use and versatility. The application’s efficacy was validated against *iglu*, an established *R* tool with a no-code web app for CGM analysis, using data from 418 participants from three studies. The unique period-specific analysis feature was demonstrated through an illustrative case study.

**Results:** *Diametrics* proved effective at replicated established CGM metrics, demonstrating high concordance with *iglu*. The platform supports a wide range of CGM devices, accommodates data in various formats, and offers extensive customization in the analysis settings. The case study highlighted *Diametrics’* ability to integrate exercise-related data with CGM readings, enabling detailed analyses of how different exercise types, intensities, and times of day impact glucose levels.

**Conclusions:** *Diametrics* is a freely available, reproducible, user-friendly, and accurate web-based tool for CGM data analysis with a unique capability to analyze data over specific time periods. With its intuitive design and open-source accessibility, *Diametrics* provides a valuable resource in diabetes research and management, empowering users of various technical levels to perform complex analyses with ease.

## Introduction

Continuous glucose monitoring (CGM) has transformed diabetes management by enabling real-time tracking of blood glucose levels and is becoming increasingly central in the management of both type 1 and type 2 diabetes (1,2). CGM devices not only allow for better daily management decisions regarding diet, medication, and lifestyle (3) but also offer researchers unprecedented data to study diabetes.

Recent advances in CGM technology have opened new research avenues focused on understanding the impact of specific events or interventions on blood glucose levels (4–13). This ability to analyze data around particular events such as meals or exercise is crucial for uncovering the nuanced effects of lifestyle factors and physiological changes on glucose control. However, the complexity of such detailed analyses often requires technical (coding) expertise, limiting access for many researchers.

Web applications (apps) offer a user-friendly solution to this challenge, requiring minimal technical skills and offering direct access through any web browser without the need for downloads. Currently, three web apps exist for CGM data analysis, offering varying degrees of complexity and functionality (14–16). These tools, however, lack the flexibility needed for in-depth examination of specific data segments, such as postprandial periods, stages of pregnancy, or exercise, thus restricting deeper insights into short-term glycemic responses around specific events.

In this paper, we introduce *Diametrics*, a novel web application designed specifically to address this need. We will validate its performance against *iglu* (14), an established *R* package for CGM data analysis that includes a web application interface. Finally, through a detailed case study, we will demonstrate how *Diametrics* can be used to analyze CGM data over specific time windows in order to closely examine glucose fluctuations around recorded events.

## Methods

### Software

*Diametrics* was developed using *Python 3.9*. The application’s architecture is designed using *Dash 2.7* (17) to be user-friendly, ensuring ease of navigation and interaction for users with varying levels of technical expertise. *Diametrics* is available as a free, open-source tool at diametrics.org. All code is available in a GitHub repository at <u>github.com/cafoala/diametrics-webapp</u>. *Diametrics* is also available as a Python package (18).

### Validation

In our validation process, we utilized data from three separate studies, totalling 418 participants. The Mobile Health Biometrics to Enhance Exercise and Physical Activity Adherence in Type 2 Diabetes (MOTIVATE-T2D) study (19) included 118 individuals with type 2 diabetes from the UK and Canada, who were provided with the FreeStyle Libre Pro CGM system and participated in a prescribed exercise program. We used two weeks of baseline CGM data for each participant. Additionally, we included two studies conducted by the Jaeb Center for Health Research (JCHR): the Type 1 Diabetes EXercise Initiative (T1-DEXI) Study (11), and the Type 1 Diabetes EXercise Initiative Pediatric (T1-DEXIP) study (12), which explore exercise in adults and adolescents with type 1 diabetes, respectively. Participants wore a Dexcom G6 CGM system for 28 days and 10 days, respectively. We randomly selected 150 participants from each of the two JCHR studies.

To validate our tool, we calculated 13 different metrics (Table 1) using both *Diametrics* and *iglu’s R* package (14). We chose the *iglu* R package as the benchmark for validation due to its thorough validation, comprehensive documentation, and its wide recognition in the field as evidenced by numerous citations. The metrics include all those recommended in the American Diabetes Association (ADA)’s International Consensus on the Use of Continuous Glucose Monitoring (20) and the Clinical Targets for Continuous Glucose Monitoring Data Interpretation (21), with the exception of the number of hypo- and hyper-glycemic events. We found the definition of how to calculate these events to be ambiguous in both the International Consensus (20) and the Clinical Targets (21), a limitation also noted by the iglu team (22), and thus we chose to exclude them.

**Table 1.** Metrics used for validation of the Diametrics web application. This table offers a clear description of the 13 metrics tested in the validation and provides the corresponding mathematical formulas used for their calculation.

| Metric | Description | Calculation method |
| --- | --- | --- |
| **Data Sufficiency** | Percentage of data available for analysis. | $\frac{\text{Number of valid glucose readings}}{\text{Total possible readings}} \times 100$ |
| **Percentage Time in Normal Range** | Percentage of readings where glucose levels are in normal range (70-180 mg/dL). | $\frac{\text{Readings} \in (70, 180) \text{ mg/dL}}{\text{Number of readings}} \times 100$ |
| **Percentage Time in Hypoglycemia** | Percentage of readings where glucose levels are in hypoglycemia (70 mg/dL). | $\frac{\text{Readings} < 70 \text{ mg/dL}}{\text{Number of readings}} \times 100$ |
| **Percentage Time in Hyperglycemia** | Percentage of readings where glucose levels are in hyperglycemia (>180 mg/dL). | $\frac{\text{Readings} > 180 \text{ mg/dL}}{\text{Number of readings}} \times 100$ |
| **Average Glucose** | Mean of all glucose readings. | $\frac{\Sigma \text{Glucose readings}}{\text{Number of readings}}$ |
| **Standard Deviation (SD)** | Measure of the glucose reading variability. | $\sqrt{\frac{\Sigma (\text{Glucose reading} - \text{Average glucose})^2}{n - 1}}$ |
| **Coefficient of Variation (CV)** | Relative measure of glucose reading variability. | $\frac{\text{Standard deviation}}{\text{Average glucose}} \times 100$ |
| **Estimated hbA1c (eA1c)** | Estimated average glucose over the past 2-3 months. | $\frac{28.7 \times \text{Average glucose (mg/dL)} + 46.7}{1.59}$ |
| **Low Blood Glucose Index (LBGI)** | Measure of frequency and extent of hypoglycemia | $\sum_{i=1}^n \left( 1.1 - \left( \frac{\text{glucose}_i}{100} \right)^2 \right)$for readings <70 mg/dL |
| **High Blood Glucose Index (HBGI)** | Measure of frequency and extent of hyperglycemia | $\sum_{i=1}^n \left( \left( \frac{\text{glucose}_i}{100} \right)^2 - 1 \right)$for readings >180 mg/dL |
| **Area Under the Curve (AUC)** | Integral of glucose values over time, indicating overall exposure to glucose. | $\sum_{i=1}^{n-1} \frac{(\text{glucose}_i + \text{glucose}_{i+1})}{2} \times (\text{time}_{i+1} - \text{time}_i)$ |

We compared the metrics from each tool using Pearson correlation coefficients and Bland-Altman plots. This enabled us to quantitatively assess the degree of correlation and agreement between the two sets of results, thereby providing a robust measure of *Diametrics’* validity and reliability in analyzing CGM data across different studies and devices.

By validating *Diametrics* against *iglu*, this also demonstrates concordance between *Diametrics* and the *cgmanalysis* (23) and *CGManalyzer* (24) packages, since *iglu* has been previously validated against them (14).

### Case study

To showcase the capabilities of *Diametrics* to perform in-depth data analysis into specific time windows, we present an illustrative example. To our knowledge, the analysis undertaken in this example cannot be replicated using any other available web app without technical ability or significant data processing by the user.

In this analysis, we used data from a study conducted by Exercise in Type 1 Diabetes (EXTOD) (25). In the trial, 34 participants with type 1 diabetes were monitored for eight weeks as they trained for an endurance running event. They wore FreeStyle Libre CGMs and recorded their exercise in a diary.

For this case study, we explored glucose dynamics during exercise and in the three hours prior and the three hours post-exercise, divided into one hour windows. We also examined how intensity (light, moderate, or vigorous), type of exercise (aerobic or anaerobic/mixed [non-aerobic]), and time of day (morning, afternoon, or evening) affects glycemic control around exercise.

We prepared the data from the exercise diary in accordance with the directions on the Diametrics documentation page, advanced analysis (diametrics.org/documentation). We included the intensity, type of exercise and time of day as additional data fields in the file. We conducted the data analysis through the *Diametrics* web app to calculate the metrics of glycemic control then downloaded the output to create a table and figure independently.

## Results

### Web App Functionality

An overview of the features of *Diametrics* and a comparison to the other available web apps is available in Table 2.

**Table 2.**
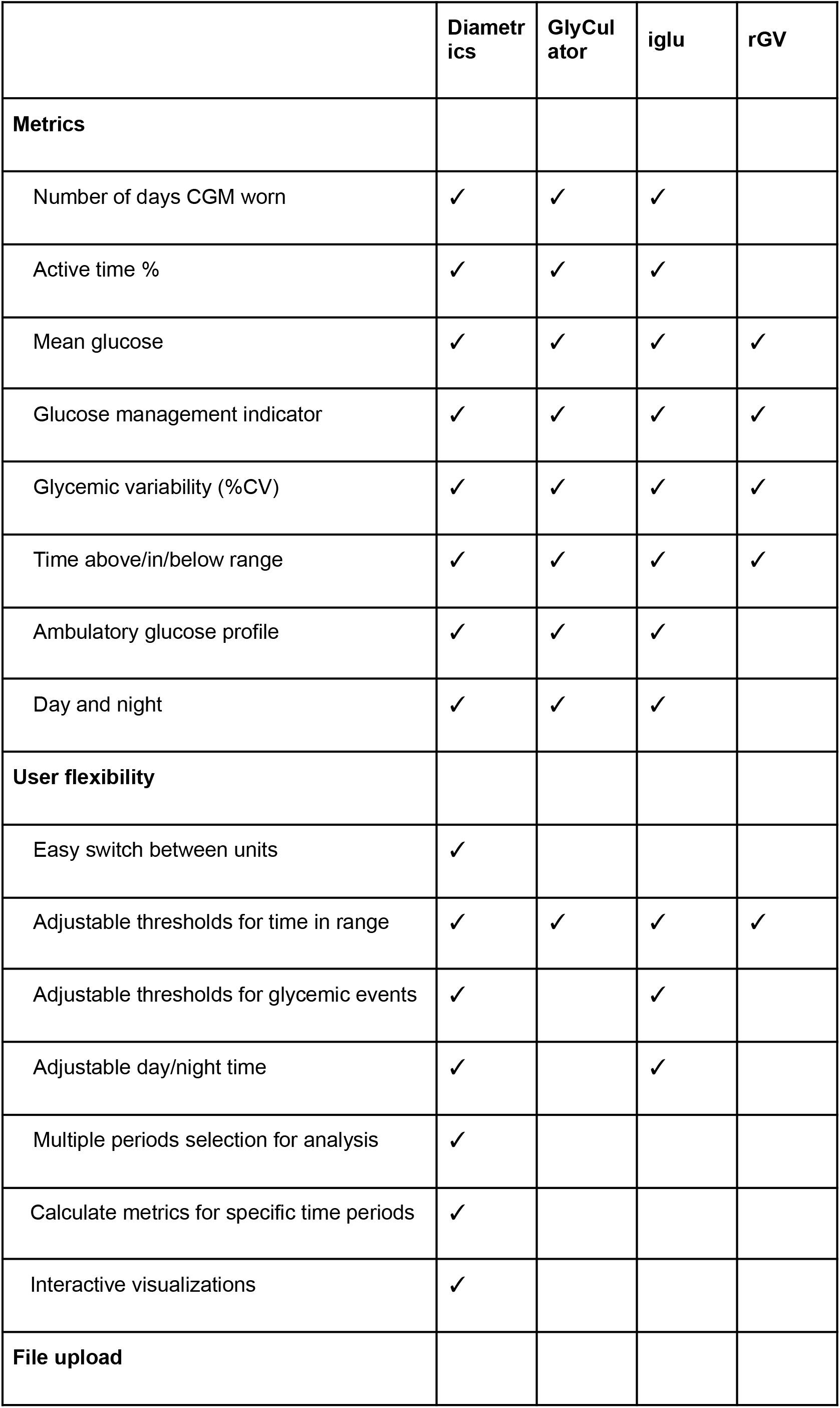
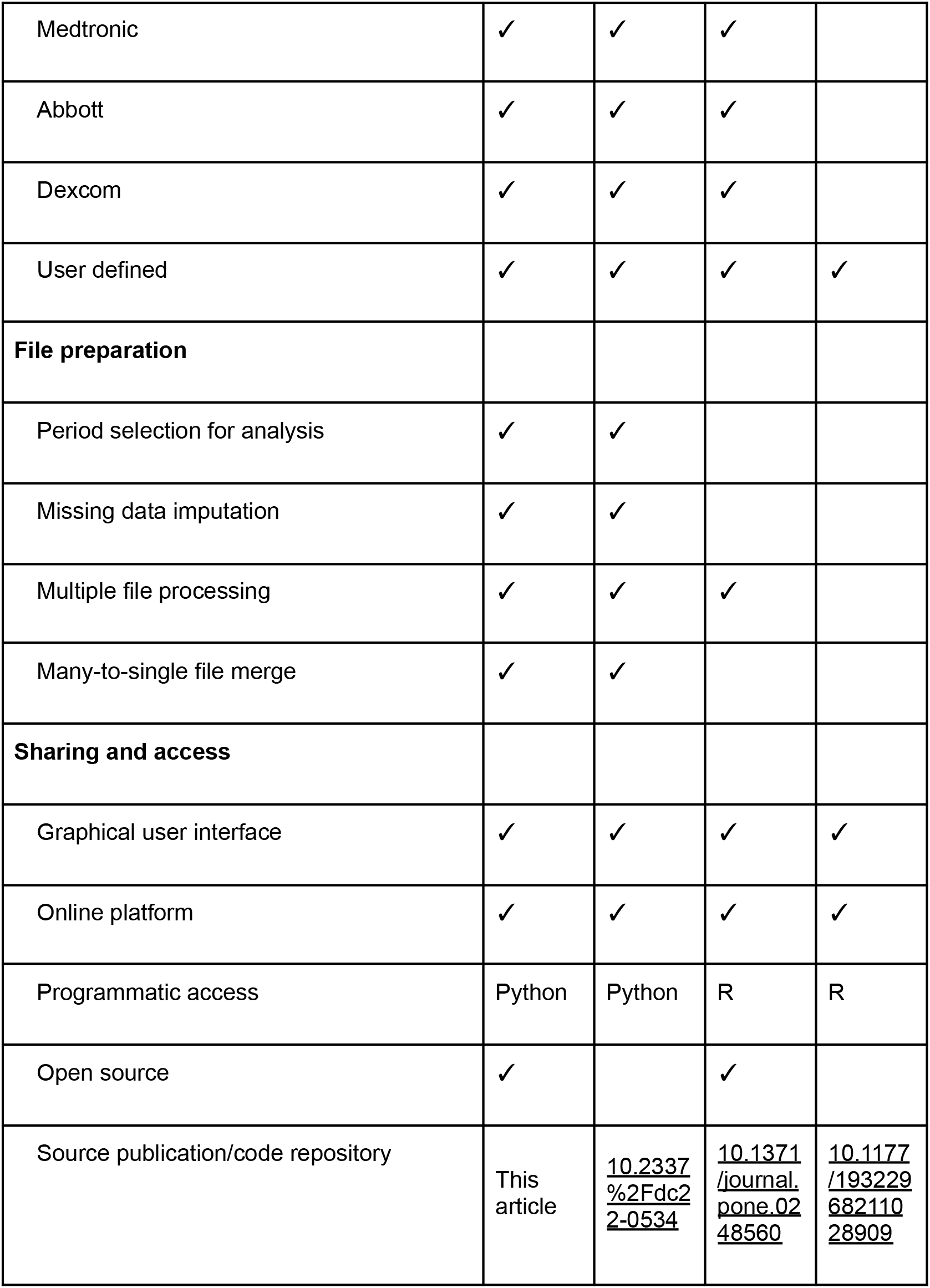
Comparison of functionality between no-code CGM analysis web applications. This table provides a detailed comparison of various functionalities across four no-code CGM analysis web applications: Diametrics, GlyCulator, iglu,and rGV. Each row represents a specific feature or capability, the presence of a checkmark (✓) indicates that the corresponding application supports the feature. The table is modified from the original version created by Chrzanowski et al. (2023).

Diametrics is introduced on its homepage, which features a brief overview video and a navigational path to the documentation section. This section includes instructional videos detailing the app’s usage, specific operational steps, frequently asked questions, and explanations of metric calculations.

When using the app there are four standard steps for analyzing the data (upload files, check data, analysis options and standard metrics) and additional steps that can be taken to visualize the data or do more advanced analysis.

#### Upload files

*Diametrics* is designed to support the upload of CGM data in various formats, including CSV, Excel, and text files, from multiple devices including Abbott, Dexcom, and Medtronic (Figure 1). Users have the flexibility to upload multiple files of any size or quantity and can directly adjust them within the application, making it suitable for handling extensive datasets. Any data uploaded by users will not be visible to developers or other users and is deleted after analysis. When uploading, the user selects the device that the data comes from, the units that glucose is measured in and the date format.

**Figure 1.**
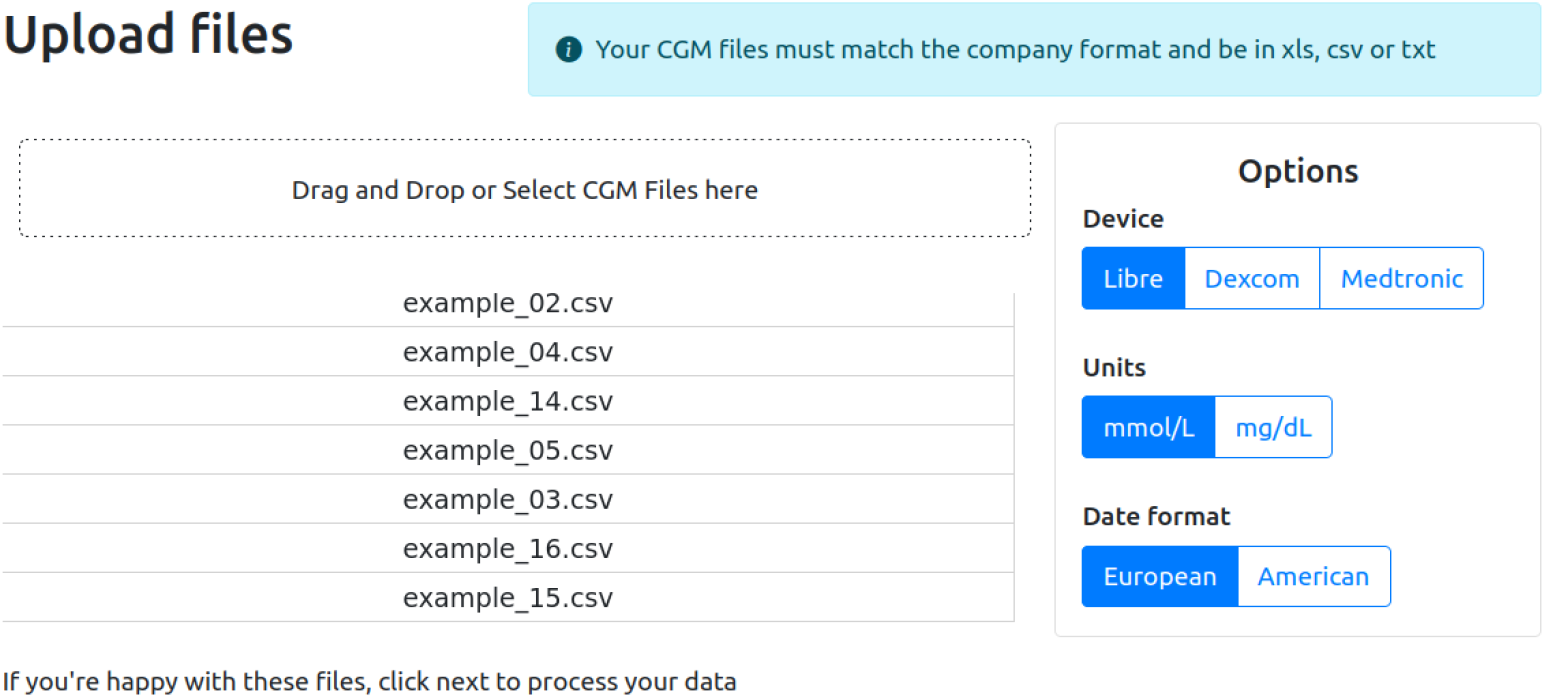
Uploading files to Diametrics. Files can be uploaded in the drag and drop box and then will be displayed under “Selected files”. The brand of CGM, the units and the date format are selected using the buttons under “Options”.

#### Check data

Once uploaded, the data is checked and displayed (Figure 2). If the data is not usable then “No” will appear in the usable column and the row will be highlighted red. The ID and start and end time of each file can be changed. If the start and end date of the file is changed, the “Days” and “Data Sufficiency (%)” columns will be updated automatically. The International consensus (20) specifies that there should be a minimum of two weeks of data with 70-80% data sufficiency. If either the number of days or the data sufficiency are below this recommendation, they will be highlighted orange to let the user know but the user can still continue with the analysis.

**Figure 2.**
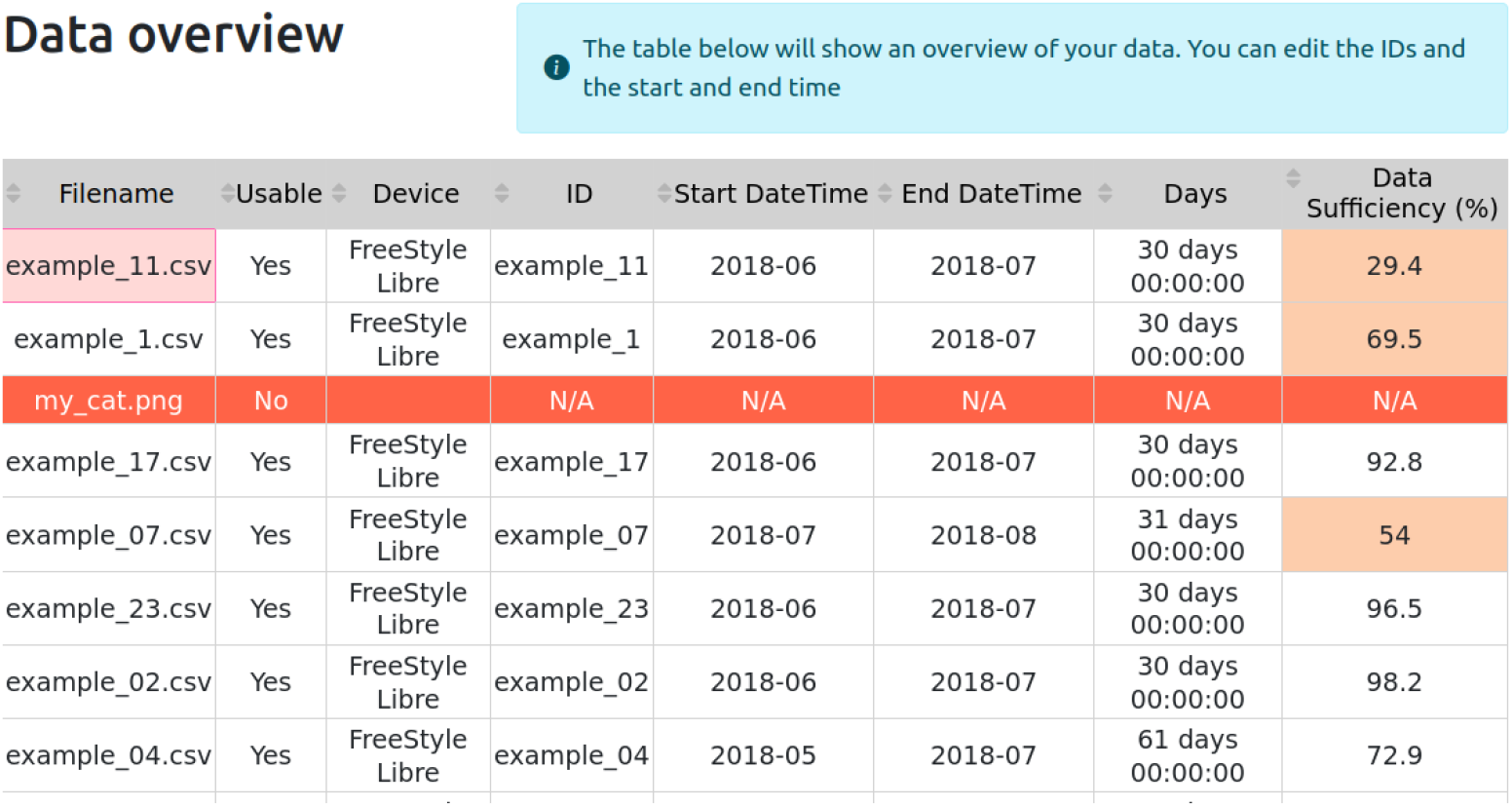
Data Validation in Diametrics. Upon uploading, the data is assessed for usability; non-usable files are marked “No” in the usable column and highlighted in red. Users can modify the ID, start, and end times for each file, automatically updating the "Days" and "Data Sufficiency (%)" columns. Entries that don’t have a minimum of two weeks of data with 70% sufficiency are highlighted in orange, though analysis can still proceed.

#### Analysis options

The application aligns with ADA’s International Consensus, offering all of the standard metrics of glycemic management. Flexible analysis options allow users to adjust the metrics to suit their specific needs (Figure 3). This includes the ability to fill gaps in data using interpolation (linear, cubic or Akima), adjust day/night time frames, set custom thresholds for time in range, and provide customizable definitions for glycemic events by changing the duration and threshold considered to be an event.

**Figure 3.**
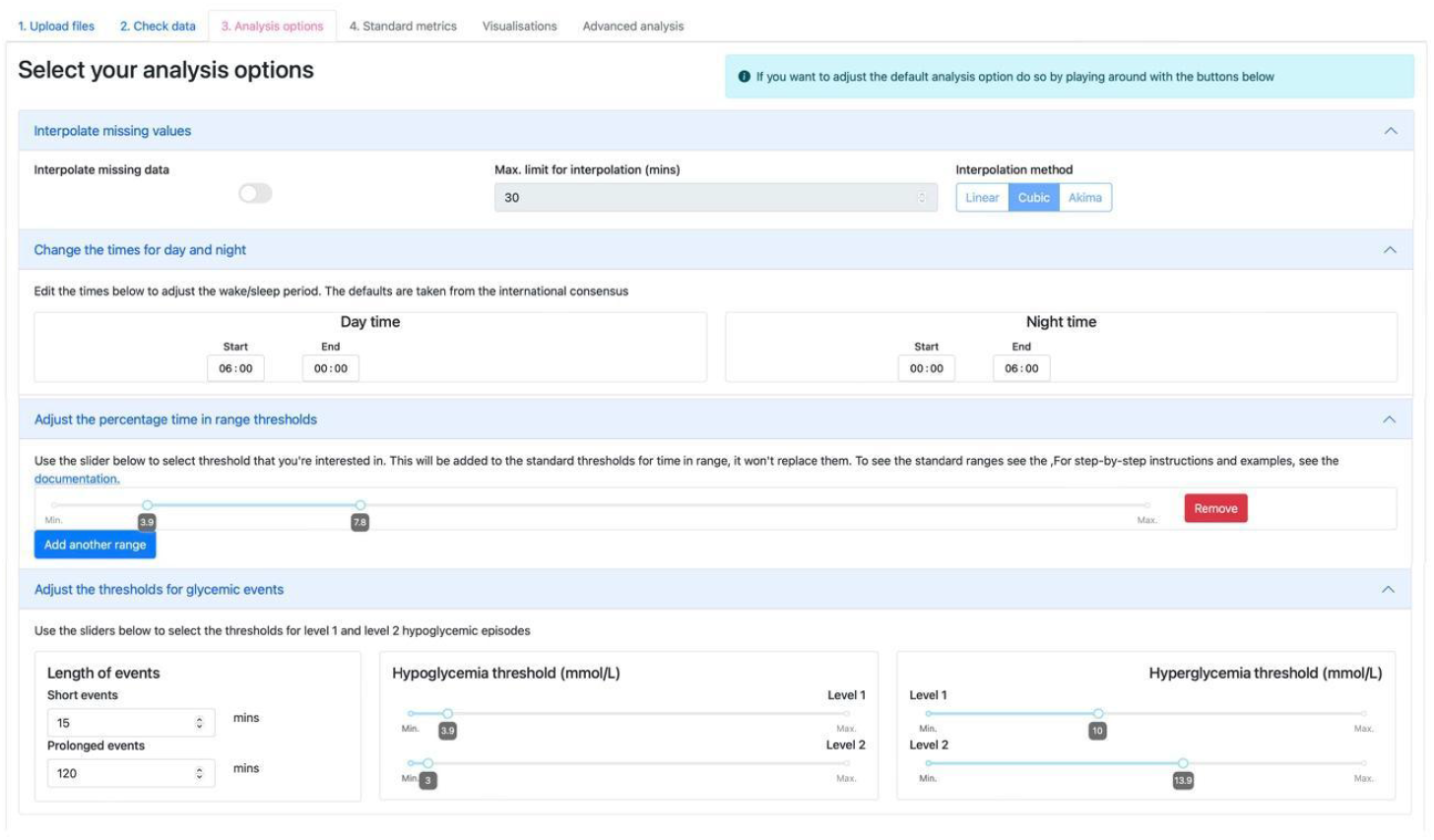
Selecting analysis options in Diametrics. Users can tailor their metrics by interpolating data gaps, altering day/night time frames, setting custom thresholds for time in range, and adjusting duration and thresholds of glycemic events.

#### Standard metrics

Within the web app, metrics are displayed in a table in which the data shown can be chosen, units switched between mmol/L and mg/dL and the metrics displayed for 24 hours or split by day or night (Figure 4).

**Figure 4.**
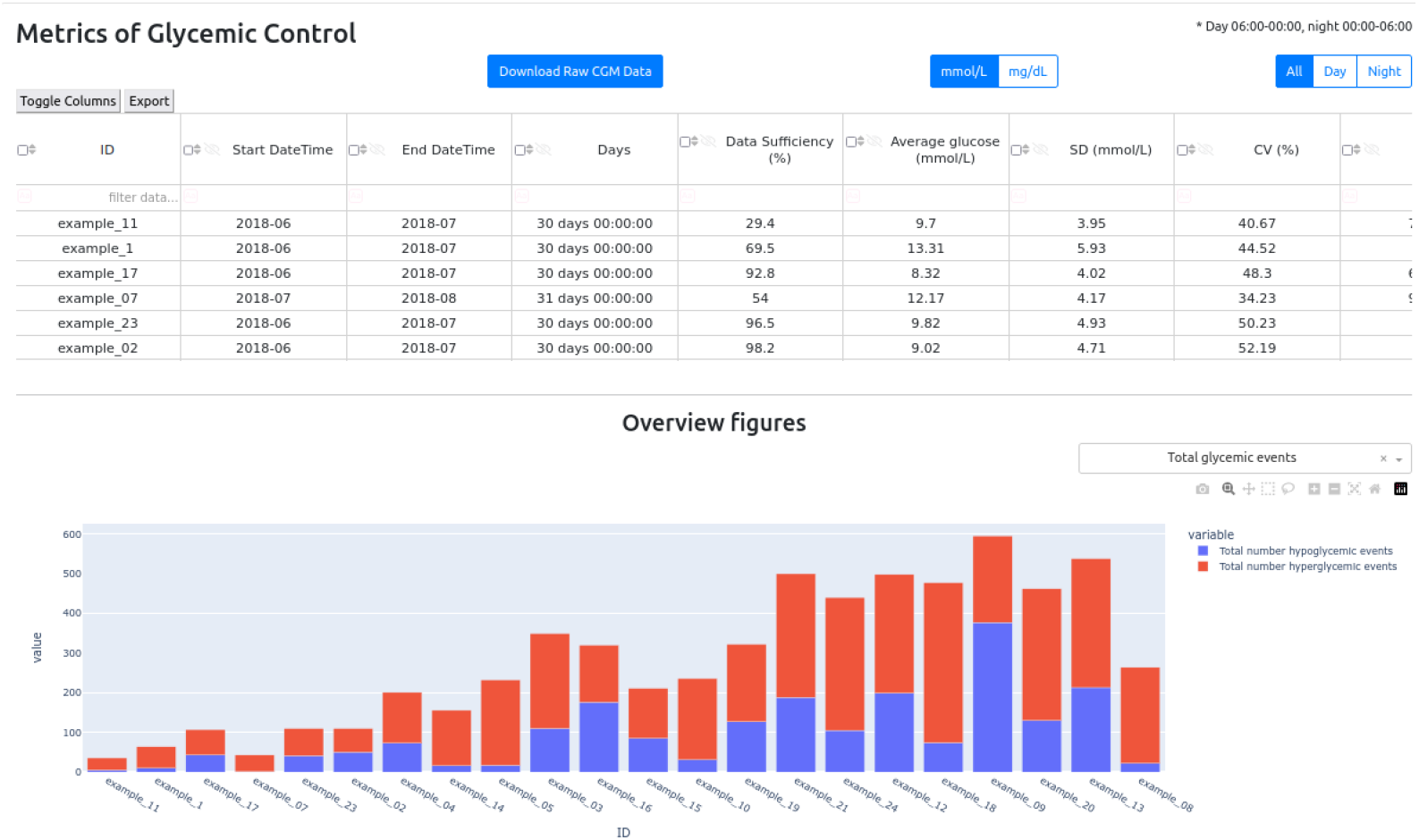
Calculating overview metrics and visualizations in Diametrics. The metrics display table within the web app allows users to select data, switch units, and choose to view metrics for a 24-hour period or split by day and night. Both tailored tables and combined CGM data can be downloaded for further analysis. The overview figures can be changed to show different metrics, they are interactive and downloadable.

Additionally, the web app allows users to customize how metrics are displayed in a table by selecting specific data to be shown, switching units between mmol/L and mg/dL, and choosing to view metrics over a 24-hour period or divided into day and night segments (Figure 4). Any user-adjusted metrics are also displayed here. Users can download their tailored tables and original combined CGM data for further analysis. Customized overview figures are automatically created to reflect the data in the table and are both interactive and downloadable.

#### Visualizations

For data visualization, *Diametrics* offers interactive graphs and charts using *Plotly* (26), which enhances data interpretation and pattern identification. These visualizations are not only informative but also downloadable and can be manipulated by users. Two example charts are shown in Figure 5, including A) ambulatory glucose profile for one participant, and B) percentage time in range for 8 participants.

**Figure 5.**
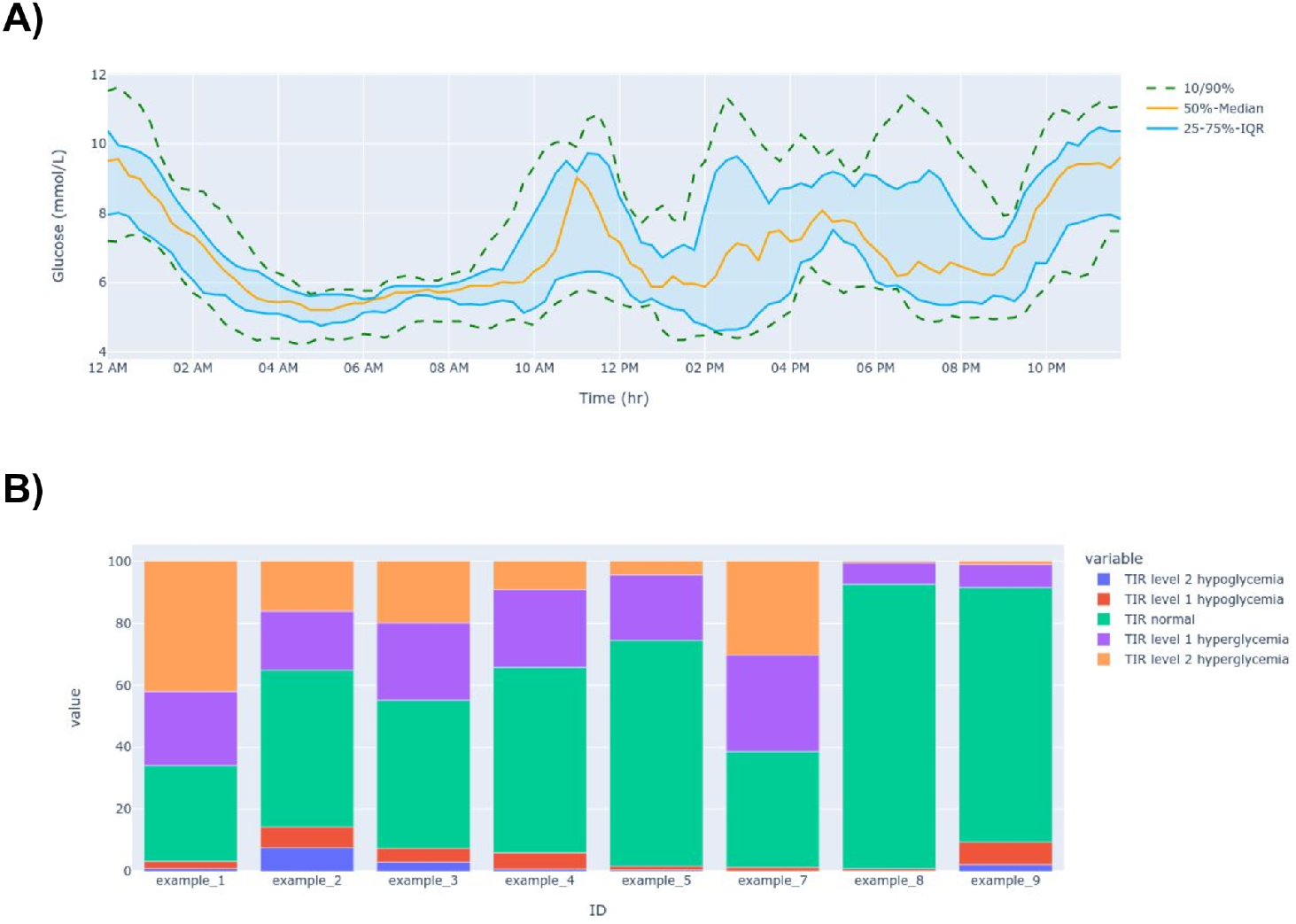
Examples of interactive figures available in Diametrics. This figure showcases two of the interactive figures accessible within the Diametrics application. A) Ambulatory Glucose Profile for One Participant; a standardized graphical representation of a patient’s glucose levels, typically over a 24-hour period, providing insights into patterns and variability of glucose control. The green dashed line represents the 10th and 90th percentiles, the orange line depicts the median glucose values, the blue shaded area represents the interquartile range. B) Percentage Time in Range for Eight Participants; detailing the distribution of time spent in various glycemic ranges for a cohort of participants. TIR level 2 hypoglycemia, percentage of glucose readings <54 mg/dL; TIR Level 1 hypoglycemia, percentage of glucose readings between 54 mg/dL and 70 mg/dL; TIR normal: percentage of glucose readings between 70 mg/dL and 180 mg/dL; TIR level 1 hyperglycemia, percentage of glucose readings between 180 mg/dL and 250 mg/dL; and TIR level 2 hyperglycemia, percentage of glucose readings >250 mg/dL.

#### Advanced analysis

A key feature of *Diametrics* is its ability to perform periodic analysis, which allows users to calculate the metrics of glycemic management for specific time frames within the CGM data, such as mealtimes or exercise sessions. Users can upload a file with detailed period information and labels, and the application will link the events to the relevant CGM data and then analyze time windows around these events, calculating both standard and user-adjusted metrics for these periods.

### Validation

In our validation analysis, *Diametrics* demonstrated a high degree of agreement when compared with *iglu’s R* package. For 12 out of the 13 evaluated metrics, including average glucose, coefficient of variation (CV), and time in range measures, this alignment is evidenced by perfect Pearson correlation coefficients of 1 across all three studies (Table 3). The only exception was found in the metric for percentage active time/data sufficiency, which still demonstrated extremely high concordance, with Pearson correlation coefficients of 1 in the MOTIVATE-T2D study but 0.999 in both the T1-DEXI and T1-DEXIP studies.

**Table 3.**
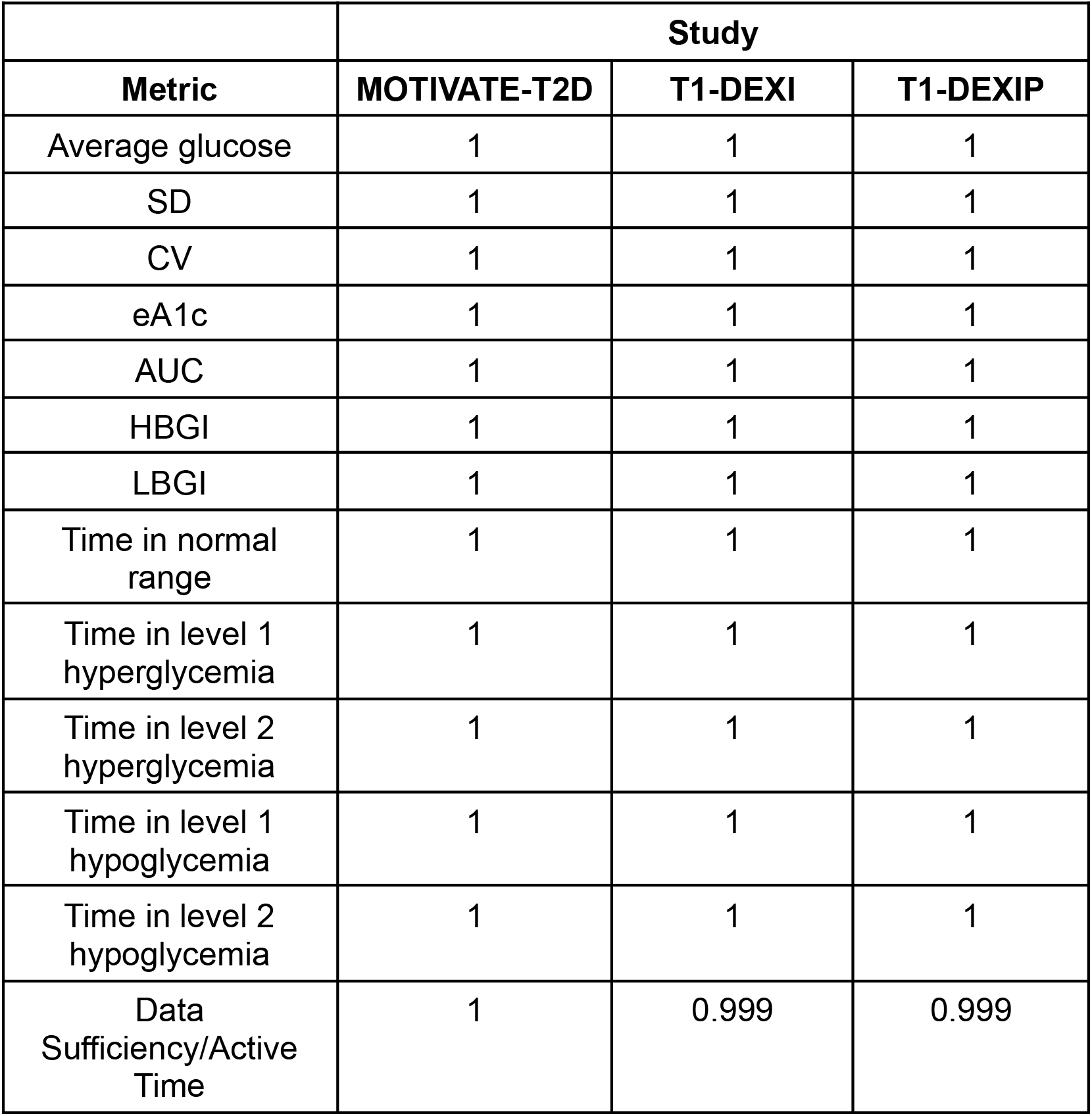
Pearson Correlation Coefficients for comparing calculations between Diametrics and IGLU for 13 common CGM metrics. This table presents the Pearson correlation coefficients for a range of metrics comparing the results from Diametrics and iglu for the MOTIVATE-T2D, T1-DEXI and T1-DEXIP studies. The values represent the correlation between the results of the two compared methodologies across these different measures, with 1 to 3 decimal places, indicating a perfect correlation. Abbreviations: Standard Deviation (SD), Coefficient of Variation (CV), estimated A1c (eA1c), Area Under the Curve (AUC), High Blood Glucose Index (HBGI), Low Blood Glucose Index (LBGI). Time in normal range, percentage of glucose readings between 70 mg/dL and 180 mg/dL; Time in level 1 hyperglycemia, percentage of glucose readings between 180 mg/dL and 250 mg/dL; Time in level 2 hyperglycemia, percentage of glucose readings >250 mg/dL; Time in level 1 hypoglycemia, percentage of glucose readings between 54 mg/dL and 70 mg/dL; Time in level 2 hypoglycemia, percentage of glucose readings <54 mg/dL. percentage of glucose readings <54 mg/dL.

The Bland-Altman plots also revealed a high level of agreement for all of the metrics. 11 of the 13 metrics demonstrated perfect agreement, with mean differences and limits of agreement of zero. AUC and data sufficiency were the only exceptions, displaying minimal disagreement. AUC exhibited a mean difference of 0.01 ± 0.10 (±1.95SD) (Figure 6A), and data sufficiency had a mean difference of -0.03 ± 0.46 (±1.95SD) (Figure 6A). It is important to emphasize that these slight differences have a negligible impact on the final results and do not significantly affect the overall validity of *Diametrics* in CGM data analysis.

**Figure 6.**
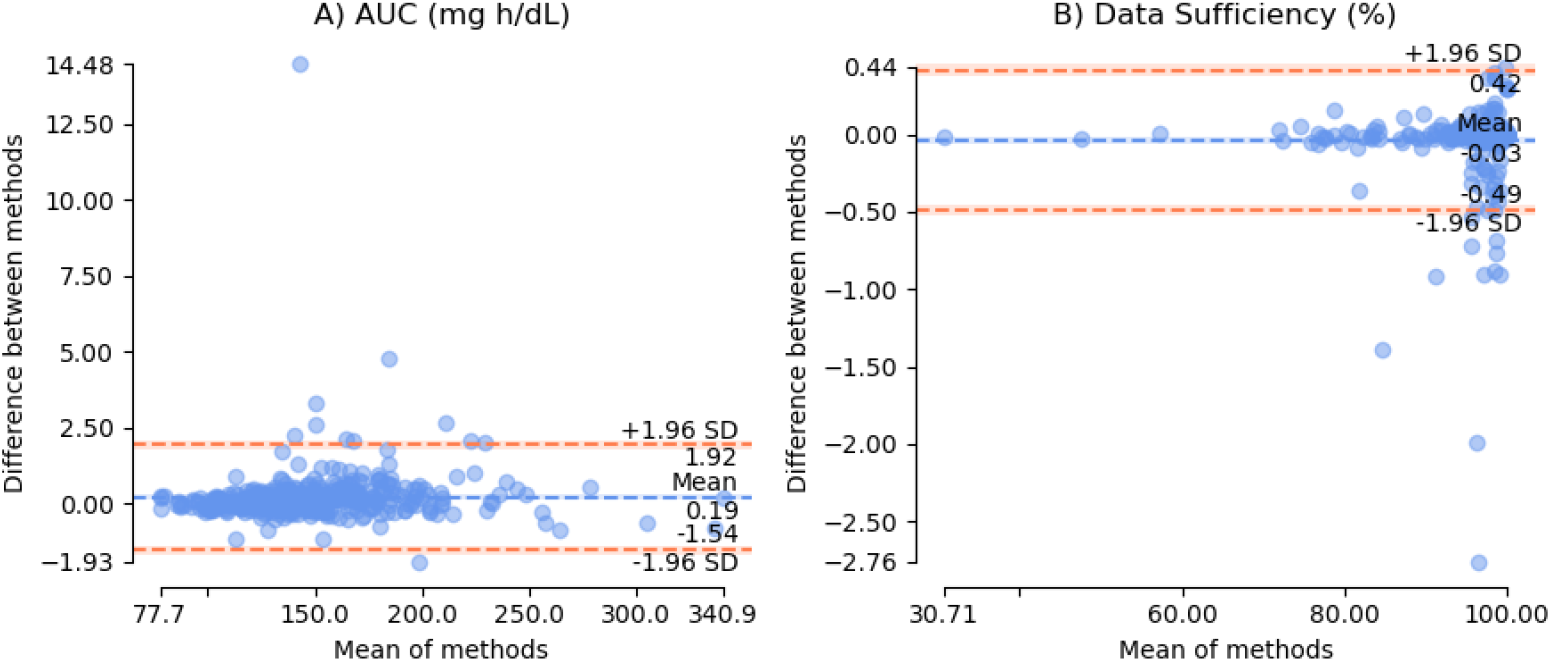
Bland-Altman Plots for Method Comparison in Glycemic Metrics. This figure presents a series of Bland-Altman plots evaluating the agreement between Diametrics and iglu methods for measuring various glycemic metrics for all three studies. Each plot illustrates the difference between the methods against the average of the two methods for a specific metric, with the metrics including: A) Area Under the Curve (AUC) measured in mg/dL, B) Data Sufficiency, representing the percentage of time with sufficient data. All other metrics demonstrated perfect agreement, with mean difference and limits of agreement of zero. The dashed lines represent the limits of agreement (mean difference ± 1.96 standard deviation [SD]), providing a visual interpretation of the consistency between the two measurement methods. Points that lie within these limits are considered to have acceptable agreement. The mean difference (bias) is depicted by the solid orange line.

### Case study

The results of the case study demonstrate the capabilities of *Diametrics* for detailed data analysis. A video of the worked case study is available at https://www.youtube.com/watch?v=bfiQRGhCLh4&t=26s.

Using the *Diametrics* web app, we were able to easily upload all CGM files simultaneously in their original downloaded format. We then set the options to match, in this case selecting FreeStyle Libre, mmol/L and European date format. Next, using the data overview section, we standardized the start and end times, which allowed for a consistent comparison of glycemic control across participants and assessed the data sufficiency of each participant to ensure the reliability of our analysis. We calculated comprehensive metrics of glycemic control for each subject, capturing the overarching patterns of glucose regulation and creating both group and individual visualizations.

Using the advanced analysis section, we uploaded an additional file containing exercise information in the compatible format. This included the start date and time of exercise, duration of exercise and three label columns identifying the time of day, type and intensity of exercise. Using only *Diametrics’* built-in functionalities, we computed the metrics for each exercise session and the respective three-hour windows pre- and post-exercise, segmented into hourly intervals. We could then download the results from *Diametrics* in order to explore variations in average glucose levels across the three-hour periods.

The observed trends in Figure 7 demonstrate a distinctive fluctuation in blood glucose levels surrounding exercise. Notably, there is an elevation in glucose concentrations leading up to the exercise period, suggestive of anticipatory action from the participants to prevent exercise-induced hypoglycemia. This is succeeded by a marked decrease post-exercise, with the most pronounced reduction occurring within the first hour following the exercise session. This pattern appears consistent across different types of exercise, intensities and times of day. There also appears to be a trend of higher glucose with higher intensity and highest glucose levels in the afternoon.

**Figure 7.**
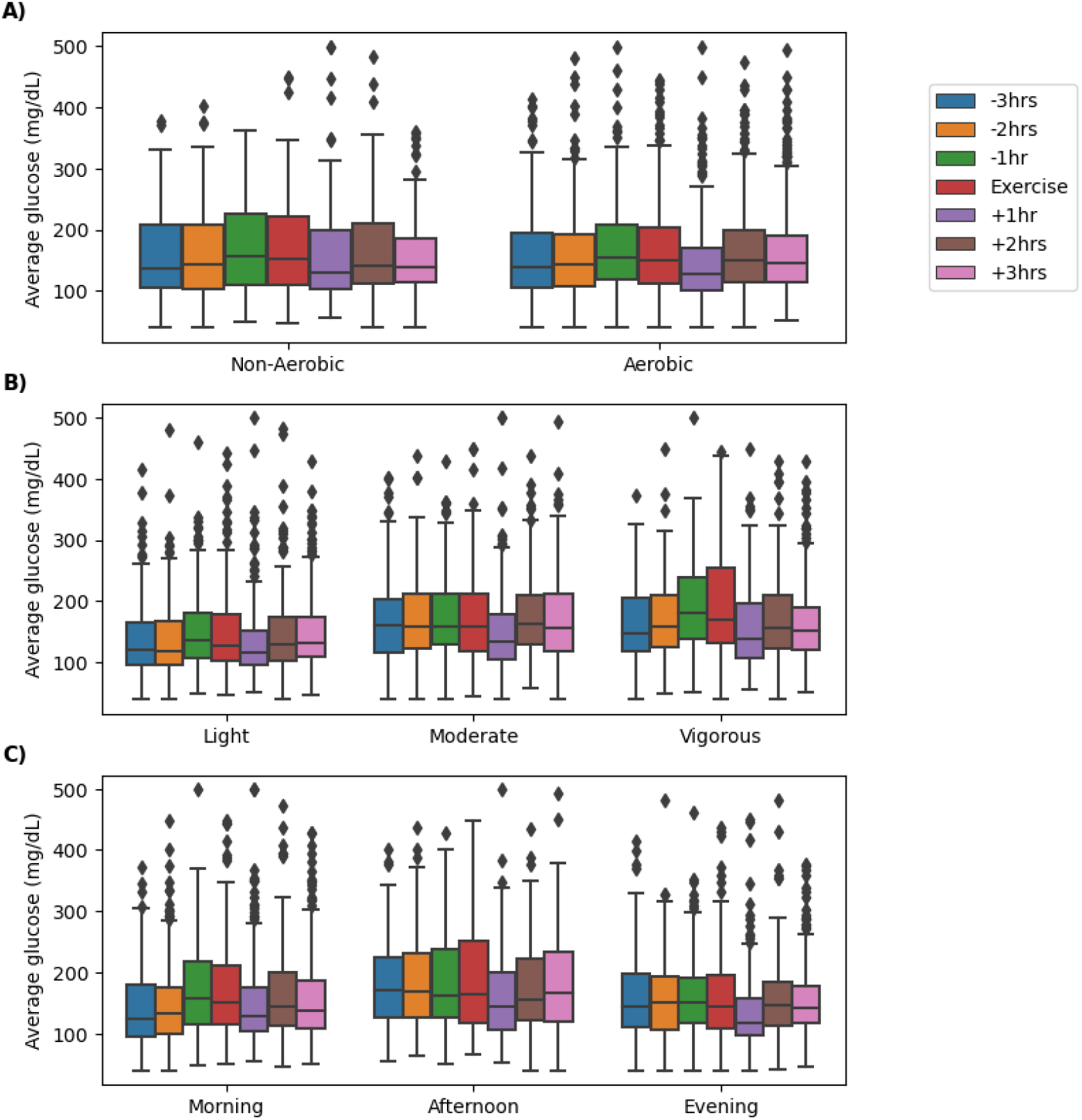
Impact of Exercise on Average Glucose Levels Before and After Exercise. Box-and-whisker plots displaying the distribution of average glucose levels (mmol/L) for 34 participants from the EXTOD 101 study for the three hours before and after an exercise bout, segmented into one-hour blocks. The periods are labeled as -3hrs (3 to 2 hours before exercise), -2hrs (2 to 1 hour before exercise), -1hr (1 hour before exercise), Exercise (during exercise), +1hr (end of exercise to 1 hour after exercise), +2hrs (1 to 2 hours after exercise), and +3hrs (2 to 3 hours after exercise). Figure A) shows these results divided into the type of exercise, B) divided by intensity, and C) by the time of day.

This real-world case study, derived using *Diametrics*, offers clear insights into exercise-induced changes in glucose control. It demonstrates how, without the need for complex coding or data analysis skills, one can uncover significant trends in glucose fluctuations associated with life events

## Discussion

In this study, we introduced, validated, and demonstrated *Diametrics*, a novel web application designed for advanced analysis of CGM data. *Diametrics* is a versatile data analysis tool offering multi-format data upload, comprehensive metrics calculation, customizable analysis options, period-specific analysis, and interactive visualizations. We validated *Diametrics* against *iglu* using CGM data from three distinct studies with a total of 418 participants, demonstrating *Diametrics’* accuracy in replicating established metrics set out in the International Consensus on the Use of Continuous Glucose Monitoring (20). Furthermore, *Diametrics’* unique feature of analyzing specific periods within CGM data was showcased through an illustrative case study, highlighting its potential to provide detailed insights into glycemic control during specific events, in this case exercise.

In comparison with existing no-code CGM analysis tools, *Diametrics* offers several advantages. As shown in Table 2, *Diametrics* provides additional features including easy unit switching, adjustable thresholds for time in range, and customizable definitions for glycemic events. Unlike most other CGM tools, it allows for the analysis of multiple periods within the CGM data, a feature particularly beneficial for detailed research studies and personalized diabetes management.

Comprehensive validation is crucial for ensuring the accuracy and reliability of digital platforms that calculate the metrics of glycemic control, given the widespread use of these metrics and their critical role in diabetes management and research. Our validation process compared metrics calculated using *Diametrics* against the *iglu* package. This package has been previously validated and as such was a robust choice as a comparator. We observed a high degree of concordance, with the only distinctions stemming from negligible variances in data sufficiency, which can likely be attributed to minor variations in the methodology employed to compute this metric. This was further corroborated by high agreement in the Bland-Altman plots. The consistency of these results across diverse studies, including MOTIVATE-T2D, T1-DEXI, and T1-DEXIP, demonstrates the robustness of *Diametrics*, particularly considering the range of participants and devices involved.

Additionally, the accessibility and adaptability of *Diametrics* are particularly significant. Metrics of glycemic control are frequently used by researchers but calculating them often requires considerable technical expertise. *Diametrics* addresses this challenge by offering a platform that simplifies these complex analyses, enabling researchers with varying levels of technical skill to engage deeply with CGM data.

The capacity to link event data, including dietary intake, physical activity, and phases of the menstrual cycle, directly to CGM data represents an advancement in diabetes management and research. This interconnectivity enables patients and healthcare providers to discern the immediate and delayed effects of lifestyle choices and phase of menstrual cycle on blood glucose levels, offering a more personalized and responsive approach to diabetes care.

From a research perspective, the amalgamation of lifestyle factors with CGM data opens new avenues for investigating the complex interplay between daily activities and diabetes management. It allows for the study of how specific interventions or behaviors correlate with glycemic outcomes in real-world settings, enhancing our understanding of effective diabetes management strategies.

*Diametrics’* open-access and open-source approach ensures it remains current and aligned with the dynamic landscape of diabetes research and management. The importance of open-source platforms lies in their ability to foster innovation and collaborative development, inviting contributions from the worldwide community.

While *Diametrics* shows potential, it’s important to acknowledge its limitations. The validation process, though thorough, was limited to specific datasets (MOTIVATE-T2D, T1-DEXI and T1-DEXIP), which may not cover all potential use cases. Future studies could expand the range of data and scenarios tested to ensure broader applicability. Additionally, as with any software tool, there is a need for continuous updates and improvements based on user feedback and technological advancements. Future versions of *Diametrics* could incorporate more advanced analytics features or integrate machine learning algorithms to predict glucose level trends, enhancing its utility in diabetes management.

## Conclusion

In conclusion, Diametrics is a freely available, accurate, and user-friendly web application for CGM data analysis with additional features not typically found in other CGM analysis tools. It enables quick and easy calculation of all the standard metrics of glycemic control for large amounts of data, provides options for user customization, allows for easy toggling between glucose measurement units, and crucially, facilitates in-depth period-specific analysis by linking glucose data with specific events such as meals or exercise. Consequently, Diametrics could help to support individuals with diabetes, healthcare providers, and researchers by enhancing their understanding of how specific events influence glucose levels and aiding in the improvement of diabetes management.

## Data Availability

This study is based on research using data from the Type 1 Diabetes EXercise Initiative (T1-DEXI) and Type 1 Diabetes EXercise Initiative Pediatric (T1-DEXIP) studies that has been made available through Vivli, Inc.
The MOTIVATE and EXTOD datasets are not currently available for public access.

## Abbreviations

ADA: American Diabetes Association
AUC: Area Under the Curve
CGM: Continuous Glucose Monitoring
EXTOD: Exercise in Type 1 Diabetes
JCHR: Jaeb Center for Health Research
MOTIVATE-T2D: Mobile Health Biometrics to Enhance Exercise and Physical Activity Adherence in Type 2 Diabetes
T1-DEXI: Type 1 Diabetes EXercise Initiative
T1-DEXIP: Type 1 Diabetes EXercise Initiative Pediatric

## Funding sources

None

## Disclosures

M.J.A. receives research funding from the National Institute for Health Research (NIHR) Applied Research Collaboration, South West Peninsula. C.L.R.’s PhD is supported by the Expanding Excellence in England (E3) fund. R.C.A. has received remuneration from Novo Nordisk, AstraZeneca, and Eli Lilly for conducting educational talks on diet and exercise for healthcare professionals. Funding and support was provided by the Royal Academy of Engineering (RAEng), London, UK, through a research fellowship in medical AI to N.V.

## Acknowledgements

We would like to thank the participants of the T1-DEXI, T1-DEXIP, EXTOD, and MOTIVATE-T2D trials who provided data on which to test Diametrics. We would also like to thank Emily Paremain and Katie Finch for technical support in building the app and Nick Jones and Joséphine Molveau for testing and feedback on the app.

This study is based on research using data from the Type 1 Diabetes EXercise Initiative (T1-DEXI) and Type 1 Diabetes EXercise Initiative Pediatric (T1-DEXIP) studies that has been made available through Vivli, Inc. Vivli has not contributed to or approved, and is not in any way responsible for, the contents of this publication.

This study/research is funded by the National Institute for Health and Care Research (NIHR) Exeter Biomedical Research Centre (BRC). The views expressed are those of the author(s) and not necessarily those of the NIHR or the Department of Health and Social Care. This research was supported by the Exeter Centre of Excellence in Diabetes (ExCEeD).

For the purpose of open access, the author has applied a Creative Commons Attribution (CC BY) license to any Author Accepted Manuscript version arising.

During the course of preparing this work, the authors used ChatGPT and Copilot for the purpose of summarizing research papers, text editing, and providing code snippets. Following the use of this tool/service, the authors formally reviewed the content for its accuracy and edited it as necessary. The authors take full responsibility for all the content of this publication.

## Notes

### Funding Statement

This study did not receive any funding

### Author Declarations

T1-DEXI & T1-DEXIP: These studies were accessed through Vivli, Inc. These studies have been conducted with ethical approval from their respective institutional review boards. Access to this data via Vivli was granted under the condition that the original ethical approvals encompass the secondary use of data for research purposes. This ensures that our use of these datasets adheres to the established ethical standards set forth by the original research protocols. MOTIVATE-T2D Study: Ethics committee/IRB of Liverpool John Moore University gave ethical approval for the original study, and this approval extends to the secondary use of de-identified data for the validation of the Diametrics app. EXTOD Study: Ethics committee/IRB of University of Birmingham gave ethical approval for the original study, and this approval extends to the secondary use of de-identified data for the validation of the Diametrics app.

## References

1. Lin R, Brown F, James S, Jones J, Ekinci E. Continuous glucose monitoring: A review of the evidence in type 1 and 2 diabetes mellitus. Diabet Med J Br Diabet Assoc. 2021 May;38(5):e14528.

2. Mian Z, Hermayer KL, Jenkins A. Continuous Glucose Monitoring: Review of an Innovation in Diabetes Management. Am J Med Sci. 2019 Nov 1;358(5):332–9.

3. Klonoff DC, Ahn D, Drincic A. Continuous glucose monitoring: A review of the technology and clinical use. Diabetes Res Clin Pract. 2017 Nov 1;133:178–92.

4. Abbie E, Francois ME, Chang CR, Barry JC, Little JP. A low-carbohydrate protein-rich bedtime snack to control fasting and nocturnal glucose in type 2 diabetes: A randomized trial. Clin Nutr. 2020 Dec 1;39(12):3601–6.

5. Gillen JB, Little JP, Punthakee Z, Tarnopolsky MA, Riddell MC, Gibala MJ. Acute high-intensity interval exercise reduces the postprandial glucose response and prevalence of hyperglycaemia in patients with type 2 diabetes. Diabetes Obes Metab. 2012;14(6):575–7.

6. Lessan N, Hannoun Z, Hasan H, Barakat MT. Glucose excursions and glycaemic control during Ramadan fasting in diabetic patients: Insights from continuous glucose monitoring (CGM). Diabetes Metab. 2015 Feb 1;41(1):28–36.

7. Majithia AR, Wiltschko AB, Zheng H, Walford GA, Nathan DM. Rate of Change of Premeal Glucose Measured by Continuous Glucose Monitoring Predicts Postmeal Glycemic Excursions in Patients With Type 1 Diabetes: Implications for Therapy. J Diabetes Sci Technol. 2018 Jan 1;12(1):76–82.

8. Nansel TR, Gellar L, McGill A. Effect of Varying Glycemic Index Meals on Blood Glucose Control Assessed With Continuous Glucose Monitoring in Youth With Type 1 Diabetes on Basal-Bolus Insulin Regimens. Diabetes Care. 2008 Apr 1;31(4):695–7.

9. Parthasarathy G, Kudva YC, Low PA, Camilleri M, Basu A, Bharucha AE. Relationship Between Gastric Emptying and Diurnal Glycemic Control in Type 1 Diabetes Mellitus: A Randomized Trial. J Clin Endocrinol Metab. 2017 Feb 1;102(2):398–406.

10. Rasmussen L, Christensen ML, Poulsen CW, Rud C, Christensen AS, Andersen JR, et al. Effect of High Versus Low Carbohydrate Intake in the Morning on Glycemic Variability and Glycemic Control Measured by Continuous Blood Glucose Monitoring in Women with Gestational Diabetes Mellitus—A Randomized Crossover Study. Nutrients. 2020 Feb;12(2):475.

11. Riddell MC, Li Z, Gal RL, Calhoun P, Jacobs PG, Clements MA, et al. Examining the Acute Glycemic Effects of Different Types of Structured Exercise Sessions in Type 1 Diabetes in a Real-World Setting: The Type 1 Diabetes and Exercise Initiative (T1DEXI). Diabetes Care. 2023 Apr 1;46(4):704–13.

12. Riddell MC, Gal RL, Bergford S, Patton SR, Clements MA, Calhoun P, et al. The Acute Effects of Real-World Physical Activity on Glycemia in Adolescents With Type 1 Diabetes: The Type 1 Diabetes Exercise Initiative Pediatric (T1DEXIP) Study. Diabetes Care. 2024 Jan 1;47(1):132–9.

13. Munan M, Oliveira CLP, Marcotte-Chénard A, Rees JL, Prado CM, Riesco E, et al. Acute and Chronic Effects of Exercise on Continuous Glucose Monitoring Outcomes in Type 2 Diabetes: A Meta-Analysis. Front Endocrinol. 2020 Aug 4;11.

14. Broll S, Urbanek J, Buchanan D, Chun E, Muschelli J, Punjabi NM, et al. Interpreting blood GLUcose data with R package iglu. PLOS ONE. 2021 Apr 1;16(4):e0248560.

15. Chrzanowski J, Grabia S, Michalak A, Wielgus A, Wykrota J, Mianowska B, et al. GlyCulator 3.0: A Fast, Easy-to-Use Analytical Tool for CGM Data Analysis, Aggregation, Center Benchmarking, and Data Sharing. Diabetes Care. 2023 Jan;46(1):e3–5.

16. Olawsky E, Zhang Y, Eberly LE, Helgeson ES, Chow LS. A New Analysis Tool for Continuous Glucose Monitor Data. J Diabetes Sci Technol. 2022 Nov 1;16(6):1496–504.

17. Hossain S. Visualization of Bioinformatics Data with Dash Bio. 2019;

18. Russon C. Diametrics: an open-source Python package for calculating common metrics of glycemic control. 2021.

19. Hesketh K, Low J, Andrews R, Jones CA, Jones H, Jung ME, et al. *Mo* bile Heal*t*h B*i*ometrics to Enhance Exercise and Physical Acti*v*ity *A*dherence in *T*yp*e* 2 Diabetes (MOTIVATE-T2D): protocol for a feasibility randomised controlled trial. BMJ Open. 2021 Nov;11(11):e052563.

20. Danne T, Nimri R, Battelino T, Bergenstal RM, Close KL, DeVries JH, et al. International Consensus on Use of Continuous Glucose Monitoring. Diabetes Care. 2017 Nov 10;40(12):1631–40.

21. Battelino T, Alexander CM, Amiel SA, Arreaza-Rubin G, Beck RW, Bergenstal RM, et al. Continuous glucose monitoring and metrics for clinical trials: an international consensus statement. Lancet Diabetes Endocrinol. 2023 Jan;11(1):42–57.

22. Broll S, Urbanek J, Buchanan D, Chun E, Muschelli J, Punjabi NM, et al. Interpreting Glucose Data from Continuous Glucose Monitors. 2024. p. 23. Available from: https://cran.r-project.org/web/packages/iglu/iglu.pdf

23. Vigers T, Chan CL, Snell-Bergeon J, Bjornstad P, Zeitler PS, Forlenza G, et al. cgmanalysis: An R package for descriptive analysis of continuous glucose monitor data. PLOS ONE. 2019 Oct 11;14(10):e0216851.

24. Zhang XD, Zhang Z, Wang D. CGManalyzer: an R package for analyzing continuous glucose monitoring studies. Bioinformatics. 2018 May 1;34(9):1609–11.

25. Bunnewell S, Thompson C, Coker P, Narendran P, Andrews R. A real-world study of people with type 1 diabetes undertaking the Swansea half marathon (Abstract). Diabetes UK Prof Conf 2021. 2021;Diabet Med 38(S1):P76.

26. Plotly Technologies Inc. Collaborative data science. 2015; Available from: https://plot.ly

